# Validation of an Afan Oromo translation of the Cutaneous Leishmaniasis Impact Questionnaire

**DOI:** 10.64898/2026.05.18.26353479

**Authors:** Debisa Eshatu Wendimu, Yohannes Hailemichael, Eyerusalem Tesfaye Beyene, Derese Bekele Daba, Sagni Chali Jira, Tedros Nigusse, Fewzia Shikur Mohammed, Shimelis Nigusie Doni, Amel Beshir Mohammed, Kibrome Tekleab, Tadele Molla, Tewodros Kaleb Kassa, Endale Hailu, Mosisa Bekele Degefa, Galana Mamo, Fikregabrail Aberra Kassa, Belachew Hailu, Teklu Cherkose, Saba Maria Lambert, Michael Marks, Stephen L. Walker, Endalamaw Gadisa, SHARP collaboration

## Abstract

**Introduction:** Cutaneous leishmaniasis (CL) is a neglected tropical disease associated with reduced health-related quality of life (HRQoL) that leads to permanent scars, anatomical damage and functional impairment. We aimed to translate, culturally adapt and validate the disease specific HRQoL measure the Cutaneous Leishmaniasis Impact Questionnaire (CLIQ) into Afan Oromo.

**Methods:** The English version of the CLIQ was translated into Afan Oromo, and culturally adapted by experts with feedback from individuals affected by CL. The finalized Afan Oromo version was then administered to adults with CL. Its psychometric properties were examined using internal reliability, inter rater reliability, construct validity, and responsiveness to change. In addition, the clinical importance difference (CID) and cut-off points for the total CLIQ score were determined.

**Results:** The Afan Oromo CLIQ demonstrated acceptable content validity, with I-CVI values ranging 0.83 to1.00. One hundred and forty-four individuals with confirmed CL with a mean age of 35.5 (±16.5) years were interviewed using the Afan-Oromo version of the CLIQ. The overall median CLIQ score was 40 (IQR=24). The median score for general impacts of CL (Cluster-1), and perceptions about health services and treatment (Cluster-2) were 32 and 9 respectively. The internal consistency (Cronbach alpha= 0.87) and inter-rater reliability (ICC=0.98) were excellent. The differences in median CLIQ scores between physicians determined CL severity classifications and between small and larger lesions were significant. The Afan Oromo CLIQ was responsive to change following treatment (P = 0.037). The CID was 9 and 7 units, using distribution and anchor methods, respectively.

**Conclusion:** The Afan Oromo CLIQ is a valid and reliable disease-specific instrument to assess HRQoL of CL affected individuals.

## 1. Introduction

Cutaneous leishmaniasis (CL) is a vector-borne neglected tropical disease caused by species of the genus *Leishmania*. CL causes scarring, functional impairment and visible damage which is stigmatising particularly when affecting the face (1,2). In Ethiopia, the estimated annual incidence of CL is 40,000 new cases which are almost exclusively due to *L. aethiopica*. It is estimated that 29 million people (32.6% of the population) live in endemic areas (3). CL has three clinical phenotypes: localized CL (LCL), mucocutaneous leishmaniasis (MCL), and diffuse CL (DCL) (4). Previous studies in Ethiopia have consistently shown reduced health-related quality of life (HRQoL) associated with CL (5–7).

Generic patient reported outcome measures (PROMs) may lack sensitivity to capture unique aspects of CL, such as its social, economic, and emotional consequences (8). The Cutaneous Leishmaniasis Impact Questionnaire (CLIQ) was developed in Brazil to address these potential limitations (9). The CLIQ has 25 items in two clusters; general impacts of CL, and perceptions about health services and treatment. Each item has a possible integer score of 0 to 4, with a total CLIQ score possible from 0 to 100. Higher scores indicate worse HRQoL (8).

In Ethiopia Afan Oromo is spoken by 44 million people (40% of the population) as their first language. It is also spoken as a first language by people in Somalia, Sudan, Tanzania, and Kenya (10,11). An estimated 9.6 million people, 34.2% of the population in the highland areas of the Oromia region, Ethiopia, largely native Afan Oromo speakers, are considered at risk of CL (3). There are currently no validated CL specific PROMs available in Afan Oromo to measure HRQoL. We aimed to translate, adapt, and validate the English language version of the CLIQ for Afan Oromo-speaking adults in Ethiopia (12).

## 2. Methods

### 2.1. Translation and Cultural Adaptation

#### Translation

A dermatologist, and a professional translator, both native Afan Oromo speakers fluent in English, independently translated the English language version of the CLIQ (9) into Afan Oromo. The translators and a third native Afan Oromo speaker, with a public health background discussed the translations and produced an agreed unified Afan Oromo version. Two different translators, fluent in Afan Oromo and English, one with an MSc in sociology and the other a general medical practitioner, and unaware of the original English version, translated the Afan Oromo version of the CLIQ back into English.

#### Cultural adaptation

A seven-member expert committee, consisting of two dermatology residents, two epidemiologists, a health economist, a nurse, and a public health researcher, all native Afan Oromo speakers, assessed the cultural appropriateness of the items and options.

#### Content validity

The seven members of the committee rated the relevance of the Afan Oromo CLIQ items using a Likert scale as “Not-relevant”, “Somewhat Relevant”, “Quite Relevant”, and “Highly relevant”. The item-related content validity index (I-CVI) and average scale level content validity index (S-CVI/AVE) and modified kappa values were calculated. Items with I-CVI higher than 0.78, S-CVI/Ave higher than 0.9, and modified kappa statistics higher than 0.4 are considered good indicators of content validity (13).

#### Face Validity

We pilot tested the Afan Oromo CLIQ with five Afan Oromo-speaking adults with active CL. The instrument was administered by a trained research assistant, a native Afan Oromo speaker. Participants were asked to provide feedback on any confusing or unclear content. A final version of the Afan Oromo CLIQ was produced following feedback from the committee and pilot testing.

### 2.2. Study setting, design and period

The study was conducted between February 2023 and June 2024 at the ALERT Comprehensive Specialized Hospital (ALERT) in Addis Ababa, Jimma Teaching and Referral Hospital, in Jimma, and Gerbi Medium Dermatologic Clinic, a private clinic, in Jimma. At ALERT the participants were enrolled in a cohort study of CL (12,14).

### 2.3. Study Participant

Participants were native Afan Oromo speaking adults (18 years of age or older) with active CL confirmed by slit-skin smear microscopy. Participants were categorised as having LCL or MCL or DCL using study definitions (12). Clinical and demographic data were recorded on standardised forms. The largest diameter of CL lesions was measured using a disposable tape measure.

### 2.4. Sample Size, and Data Collection Procedures

We wished to recruit 250 participants (10 participants per item) based on the minimum desired sample for reliability and validity study tests (15). Consecutive, individuals with CL at the study facilities were approached. Trained data collectors, native Afan Oromo speakers, administered the Afan Oromo CLIQ (Supplementary file 1).

### 2.5. Reliability and Validity Assessment

#### Reliability assessments

We administered the Afan Oromo CLIQ to participants to assess the internal consistency of the instrument. To assess inter-rater reliability, two interviewers independently interviewed participants using the Afan Oromo CLIQ at least 60 minutes apart.

#### Construct validity

The known group validity was assessed in individuals who were classified as having mild or moderate or severe CL by the treating physician, the number of lesions (single and multiple), body region affected (head and neck only, trunk and /or limbs only, and multiple region (head/neck and trunk/limbs)), lesion size (<40 mm and ≥ 40 mm), and clinical phenotype (LCL or MCL).

Convergent validity was evaluated by assessing the correlation of items with their assigned cluster and discriminant validity by the extent of items with the other cluster.

#### Responsiveness to change

The CLIQ was administered on a second occasion 90 days after enrolment to all participants who completed the Day 90 follow up visit in the cohort study at ALERT (14).

#### Clinical Importance Difference

At Day 90, we collected participant opinion on their improvement using five-category Likert scale (Better, Somewhat better, Same, Somewhat worse, and Worse) as well as the CLIQ score to assess Clinical Importance Difference (CID) defined as any change in outcome score that is considered clinically relevant from the affected individual’s perspective (16).

### 2.6. Data management and analysis

Anonymised data were entered into the secure study database REDCap version 13.10.1 (17). STATA version 17 (18) was used for data analysis. Descriptive statistics were presented using frequencies and percentages. We calculated Cronbach’s alpha to determine the internal consistency of the instrument. A high Cronbach’s alpha (0.7 or above) suggests that the questions in the test are closely related and measure the same underlying concept (19). To assess inter-rater reliability, we used the intra-class correlation coefficient (ICC). The ICC results were interpreted as poor (if below 0.50), moderate (between 0.50-0.75), good (between 0.75-0.90), or excellent (above 0.90). A good to excellent ICC indicates minimal variation in scores assigned by different raters (20). Non-parametric statistical tests (Kruskal-Wallis and Mann-Whitney U) were used for known group validity assessment. Convergent and discriminant validity was assessed using the Spearman’s rho correlation coefficient. A correlation coefficient between 0.7 and 0.9 indicates a strong positive correlation, while a value above 0.9 suggests a very strong correlation (21). A Wilcoxon paired signed-rank test was used to assess responsiveness to change. P-values < 0.05 were considered statistically significant. Receiver operating characteristic (ROC) curves was used to establish cut-off points for total CLIQ score as mild, moderate, and severe impact using physician determined clinical severity. CID was assessed using distribution and anchor-based methods, employing participant opinion at Day 90 (16). The CID was calculated using the distribution method by multiplying moderate effect size (0.5) with the standard deviation of the difference in CLIQ scores between Day 1 and Day 90. The Anchor method for calculating the CID used the difference in mean change in score of the participant opinion “better” from “no change”.

## 3. Results

### 3.1. Translation and cultural adaptation

The expert committee adapted item 4 “Has Cutaneous Leishmaniasis somehow increased your health expenses?” by adding the phrase “*ji’oota ja’a darban keessatti*”, which means “in the last six months”.

#### Content validity

All items demonstrated strong content validity. Nineteen items had I-CVI values of 1.0 and modified kappa values of 1.0. The remaining six items had I-CVI values of 0.83 and the modified kappa values of 0.82 (Supplementary file 2, Table 1). The S-CVI/Ave value was 0.96.

**Table 1.** Socio-demographic and clinical characteristics of the study participants.

| Variable |  | Frequency n (%) | Median (IQR) score |
| --- | --- | --- | --- |
| Age group (in years) | 18-27 | 66 (45.8) | 34 (22) |
|  | 28-37 | 22 (15.3) | 47.5 (20) |
|  | 38-47 | 15 (10.4) | 46 (28) |
|  | 48-57 | 25 (17.4) | 50 (27) |
|  | 58-67 | 10 (6.9) | 50 (14) |
| | $\geq 68$ | 6 (4.2) | 39 (17) |
| Sex | Male | 96 (66.7) | 39 (23) |
|  | Female | 48 (33.3) | 43 (23.5) |
| Educational status | No formal education | 40 (27.8) | 50 (16.5) |
|  | Primary | 31 (21.5) | 43 (19) |
|  | Secondary | 35 (24.3) | 39 (23) |
|  | Post-secondary | 38 (26.4) | 27 (28) |
| Department attended | IPD | 78 (54.2) | 44.5 (28) |
|  | OPD | 66 (45.8) | 38.5 (19) |
| Number of lesions | Single | 113 (78.5) | 40 (23) |
| | Multiple ($\geq 2$) | 31 (21.5) | 40 (26) |
| Size of lesion | $< 40$ mm | 86 (60.1) | 38 (24) |
| | $\geq 40$ mm | 57 (39.9) | 49 (24) |
| Location of lesion | Head and neck only | 115 (79.9) | 40 (26) |
|  | Limbs and/or Trunk only | 20 (13.9) | 43.5 (18.5) |
|  | Multiple body regions | 9 (6.2) | 47 (18) |
| Clinical phenotype | LCL | 102 (70.8) | 39 (23) |
|  | MCL | 40 (27.8) | 44.5 (24) |
|  | DCL | 2 (1.4) | 53 (44) |
| Physician determined clinical severity | Mild | 54 (37.5) | 36.5 (24) |
|  | Moderate | 65 (45.1) | 44 (24) |
|  | Severe | 25 (17.4) | 51 (26) |

IPD: Inpatient department, OPD: Outpatient Department, mm: millimetre

#### Pre-final testing and debriefing

Following feedback from the pilot testing with five individuals with CL, “*Not Applicable*” was added as a response option for Item 14 (“Have you ever had difficulty during sexual intercourse because of the skin wound(s)?”. The “Not Applicable” response scores zero.

### 3.2. Participant Characteristics

One hundred and forty-four participants were recruited. The mean age was 35.5 years (SD ±16.4), and 96 (66.6%) were male. Sixty-six (45.8%) were between 18 and 27 years of age. The majority of participants had LCL (102, 70.8%), 40 (27.8%) had MCL, and two (1.4%) had DCL. Most participants (115, 79.8%) had a lesion on the head and neck region. Fifty-four (37.5%) were classified by the physician who assessed them as having mild CL, 65 (45.1%) moderate, and 25 (17.4%) severe (**Table 1**).

The median CLIQ score was 40 (IQR= 24), ranging from 4 to 77. The median score for Cluster 1 was 32, and nine for Cluster 2.

### 3.3. Validity and reliability assessment

#### Reliability Assessment

The overall Cronbach alpha score for the Afan Oromo CLIQ was 0.87. The Cronbach alpha ranged between 0.85 to 0.87 if each of the 25 items was deleted (**Table 2**).

The overall inter-rater agreement in 45 participants was excellent with ICC of 0.98 (CI: 0.96, 0.99). ICC values ranged from 0.49 to 0.98, with Item 23 (“How often have you relied on health services to provide you with supplies or to help changing the wound bandages?”) having the lowest agreement (Supplementary file 2, Table 2). The Bland-Altman plot (Figure 1.) demonstrates strong inter-observer agreement, with all but three measurements falling within the limits of agreement. The outliers were distributed evenly, indicating minimal variation and high consistency in scoring between raters.

**Table 2.** Cronbach’s alpha of the Afan Oromo CLIQ when item removed.

| Items | Cronbach's alpha if item deleted |
| --- | --- |
| Item 1 (Has CL affected your overall well-being?) | 0.8691 |
| Item 2 (Has CL interfered with your physical activities?) | 0.8626 |
| Item 3 (Has CL somehow affected your ability to work (or study) | 0.8629 |
| Item 4 (Has CL somehow increased your health expenses?) | 0.8689 |
| Item 5 (Do you consider that CL has financially damaged you family's budget?) | 0.8684 |
| Item 6 (Do you feel isolated from others since you got CL?) | 0.8610 |
| Item 7 (Have you suffered thinking that your appearance is different from people who don't have wounds on their skin?) | 0.8629 |
| Item 8 (Have you had difficulty walking, changing clothes or bathing because of the wound (s) on your skin?) | 0.8602 |
| Item 9 (Have you felt pain, burning, itching or discomfort at the site of the skin wound (s)?) | 0.8597 |
| Item 10 (Have you ever felt nervous, sad or scared because of CL?) | 0.8585 |
| Item 11 (Have you ever felt guilty or insecure about CL?) | 0.8582 |
| Item 12 (Have you ever felt embarrassed because of the skin wound(s)?) | 0.8586 |
| Item 13 (Have you ever missed work (or school) because of CL?) | 0.8582 |
| Item 14 (Have you ever had difficulty during sexual intercourse because of the skin wound(s)?) | 0.8677 |
| Item 15 (How often have you had to pay someone to replace you in work or home activities so you could go get health service?) | 0.8696 |
| Item 16 (Did you have to change the style of dressing because of other people's prejudices about their skin wounds?) | 0.8602 |
| Item 17 (How often have you avoided social activities with groups of people because of CL?) | 0.8584 |
| Item 18 (How often do you depend on someone else to accompany you to your medical appointments to treat CL?) | 0.8689 |
| Item 19 (What do you think about the medication you used to treat CL?) | 0.8801 |
| Item 20 (What did you think about how you were welcomed by the health services when you were seeking diagnosis of CL?) | 0.8735 |
| Item 21 (What did you think about how you were welcomed by the health services when you were seeking treatment for CL?) | 0.8749 |
| Item 22 (How often have you felt sick because of the medications you took to treat CL?) | 0.8734 |
| Item 23 (How often have you relied on health services to provide you with supplies or to help changing the wound bandages?) | 0.8746 |
| Item 24 (How much do you care about the need to seek health services for the treatment of CL?) | 0.8694 |
| Item 25 (How long has it taken to get the tests done, medical appointments or hospitalizations related to CL?) | 0.8716 |
| Total CLIQ Score | Overall= 0.8709 |

**Figure 1.**
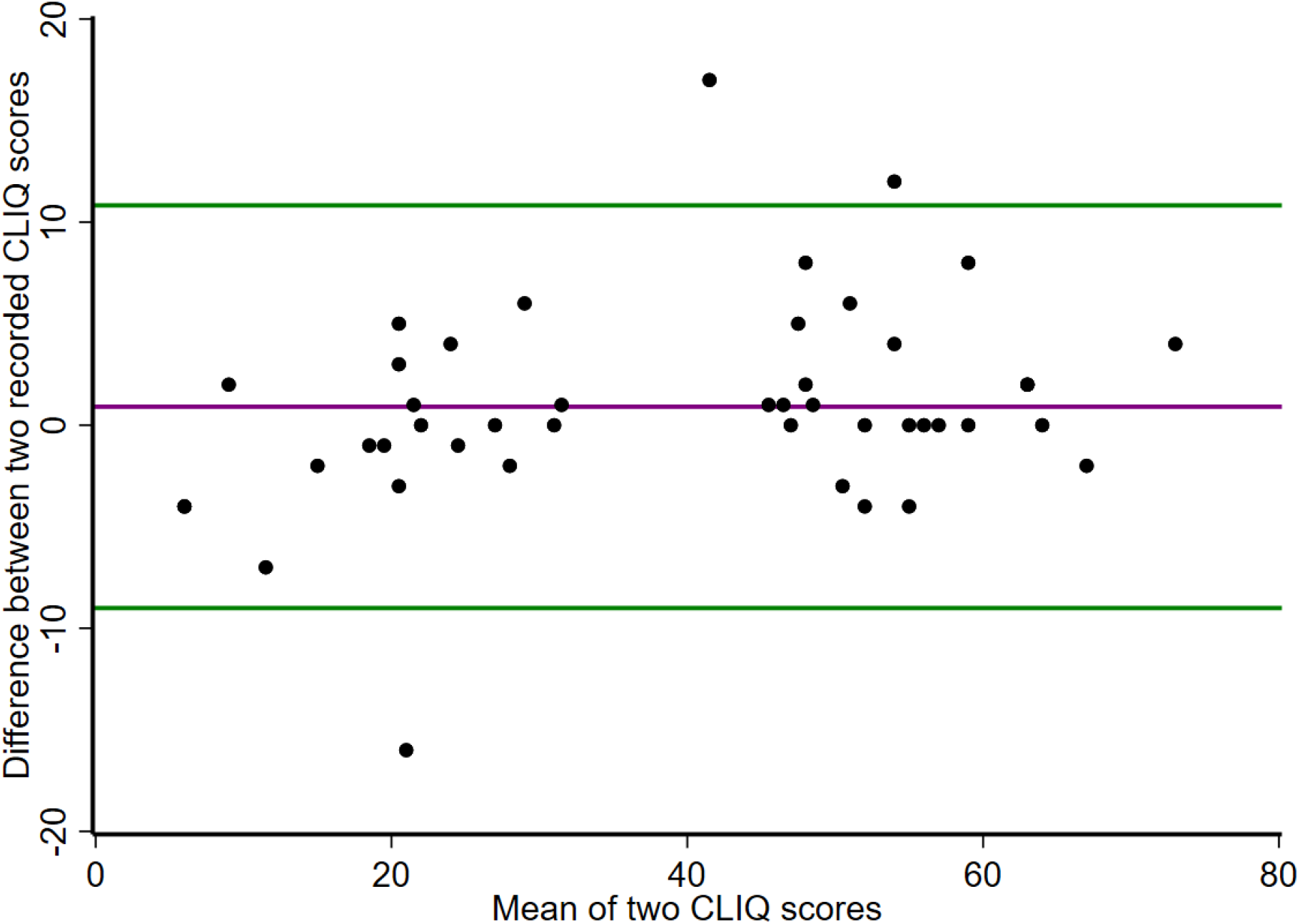
Bland-Altman plot of the difference between the Afan Oromo CLIQ scores of the two raters

#### Construct Validity

The total Afan Oromo CLIQ scores between different CL severity groups were statistically significant (p-value= 0.0061). Post-hoc pairwise comparisons, showed statistically significant differences between ‘Mild’ and ‘Moderate’ (p-value = 0.0246), and ‘Mild’ and ‘Severe’ groups (p-value = 0.0054). However, there was no significant difference between the ‘Moderate’ and ‘Severe’ groups (**Figure 2**).

**Figure 2.**
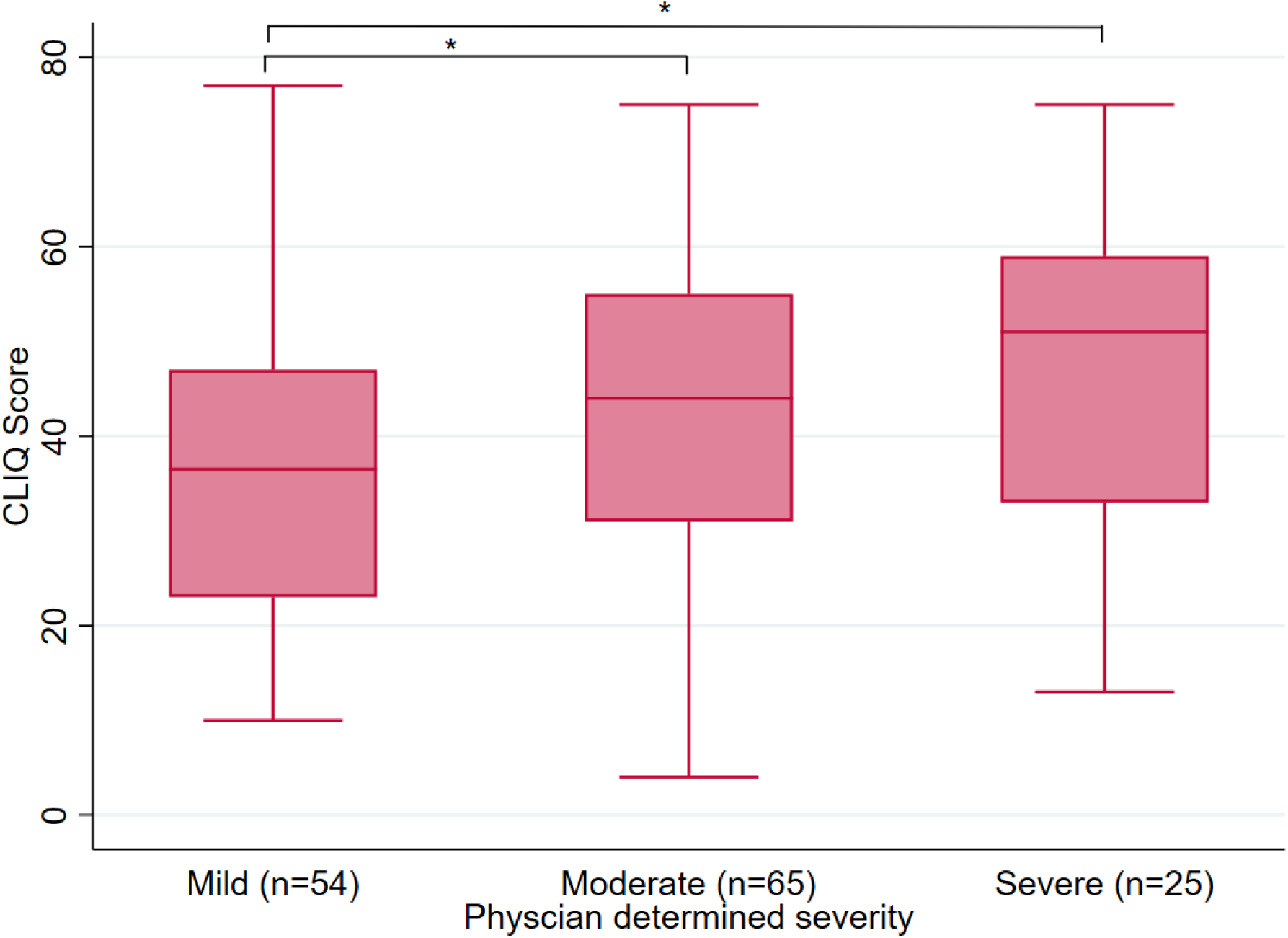
Box-plot of the CLIQ score by physician-determined severity (Note: * significant at p value <0.05).

Participants with larger lesions ≥ 40 mm longest diameter) had significantly higher Afan Oromo CLIQ scores than those with smaller lesions (<40 mm longest diameter) (p-value= 0.0027) (Supplementary file 2, Table 3).

The results for convergent and divergent validity (Table 3), showed that items had low to high correlation (0.37 to 0.78) with their assigned cluster and negligible to low correlation with the other cluster (-0.01 to 0.33).

**Table 3.**
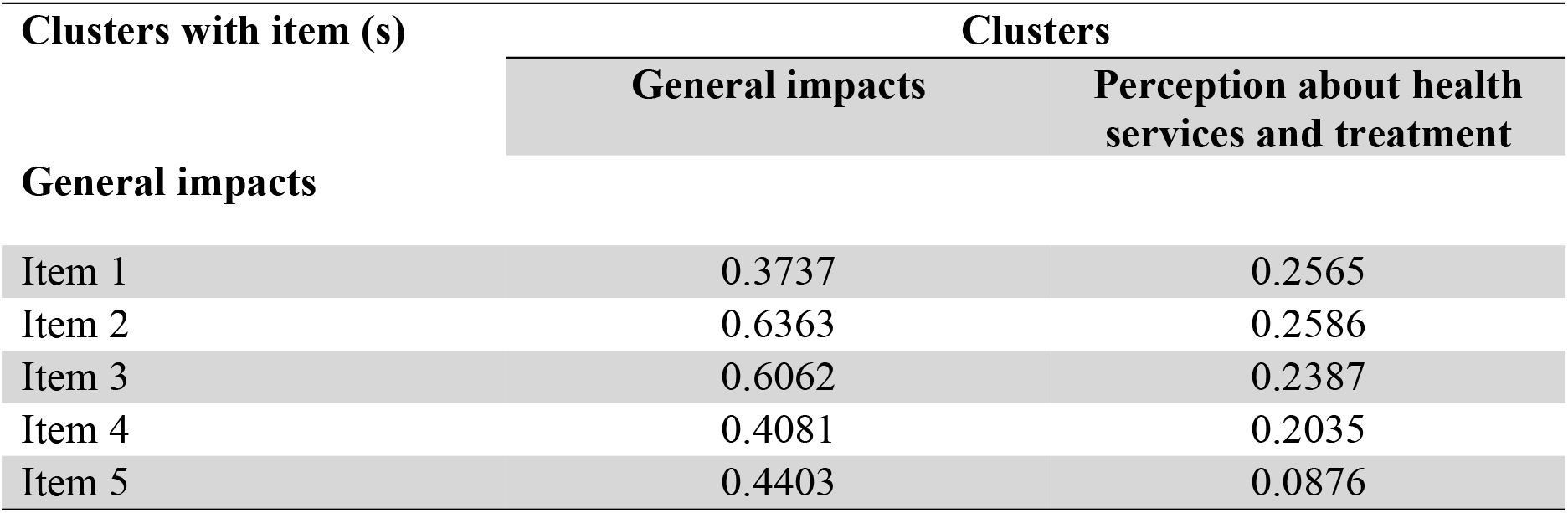
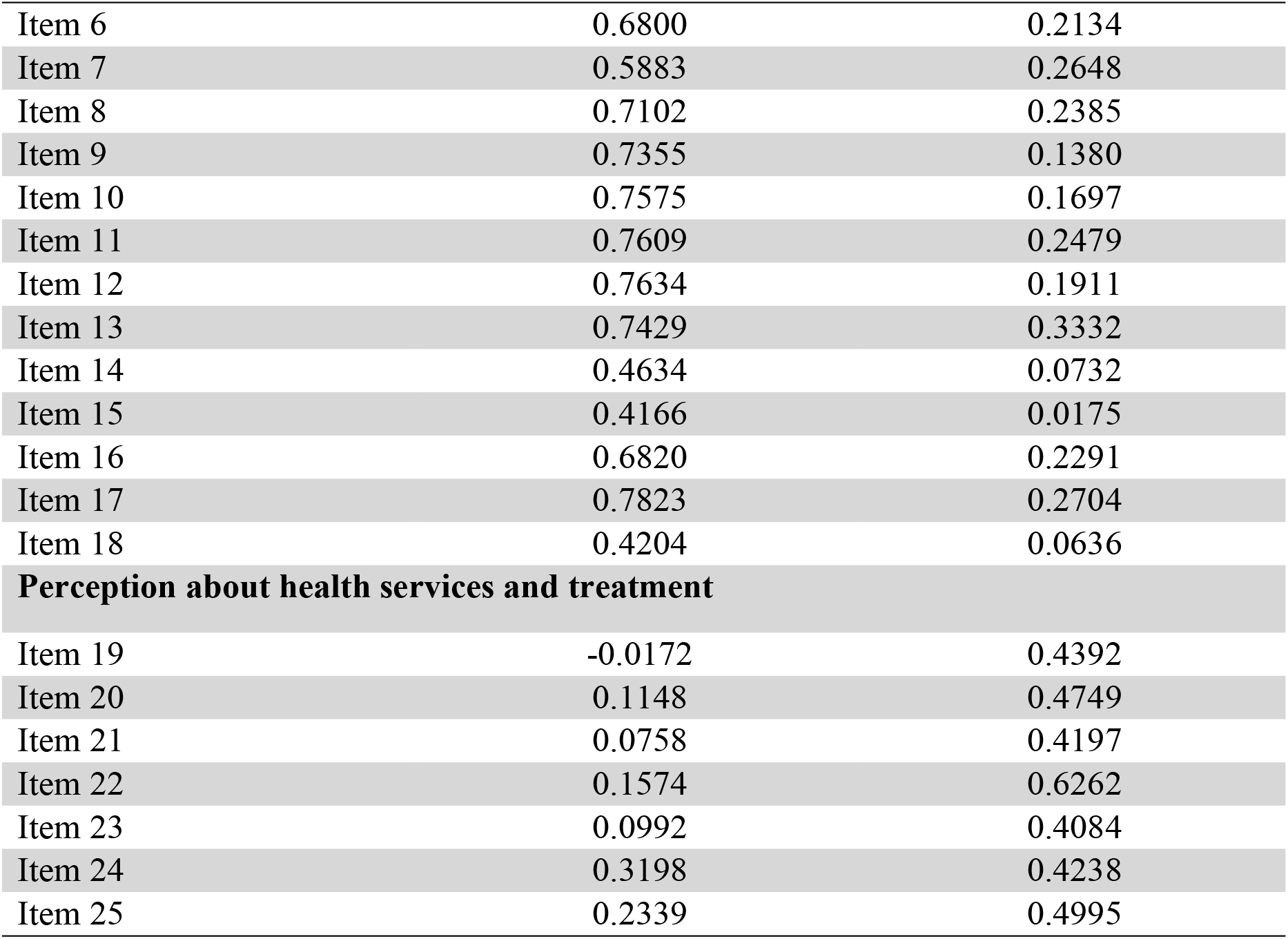
Convergent and discriminant validity of CLIQ (n=144)

| Clusters with item (s) | Clusters |  |
| --- | --- | --- |
|  | General impacts | Perception about health services and treatment |
| **General impacts** |  |  |
| Item 1 | 0.3737 | 0.2565 |
| Item 2 | 0.6363 | 0.2586 |
| Item 3 | 0.6062 | 0.2387 |
| Item 4 | 0.4081 | 0.2035 |
| Item 5 | 0.4403 | 0.0876 |
| Item 6 | 0.6800 | 0.2134 |
| Item 7 | 0.5883 | 0.2648 |
| Item 8 | 0.7102 | 0.2385 |
| Item 9 | 0.7355 | 0.1380 |
| Item 10 | 0.7575 | 0.1697 |
| Item 11 | 0.7609 | 0.2479 |
| Item 12 | 0.7634 | 0.1911 |
| Item 13 | 0.7429 | 0.3332 |
| Item 14 | 0.4634 | 0.0732 |
| Item 15 | 0.4166 | 0.0175 |
| Item 16 | 0.6820 | 0.2291 |
| Item 17 | 0.7823 | 0.2704 |
| Item 18 | 0.4204 | 0.0636 |
| **Perception about health services and treatment** |  |  |
| Item 19 | -0.0172 | 0.4392 |
| Item 20 | 0.1148 | 0.4749 |
| Item 21 | 0.0758 | 0.4197 |
| Item 22 | 0.1574 | 0.6262 |
| Item 23 | 0.0992 | 0.4084 |
| Item 24 | 0.3198 | 0.4238 |
| Item 25 | 0.2339 | 0.4995 |

Using ROC curves for physician determined clinical severity, we determined the following cut-off points for the CLIQ total score: a mild impact is characterized by a score of less than 50, a moderate impact by a score between 50 and 54, and a severe impact by a score greater than 54. The area under the curve (AUC) for distinguishing mild and moderate categories is 0.63, with a sensitivity of 38.46% and specificity of 87.0% (Figure 3a). For distinguishing moderate and severe categories, the AUC is 0.58, with a sensitivity of 44.0% and specificity of 72.3% (Figure 3b).

**Figure 3a.**
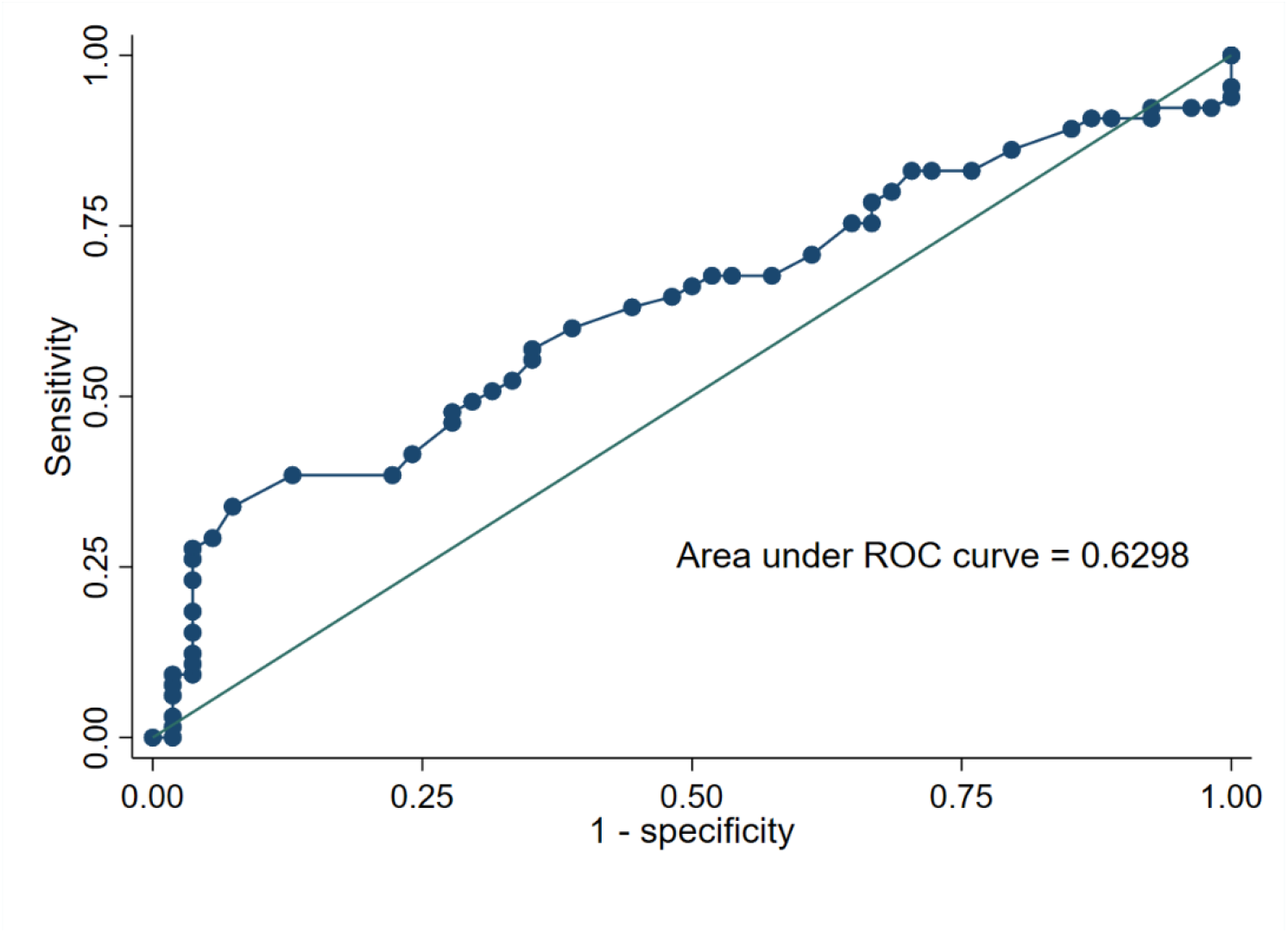
ROC curve for determining the CLIQ total score cut-off point distinguishing mild and moderate impact categories.

**Figure 3b.**
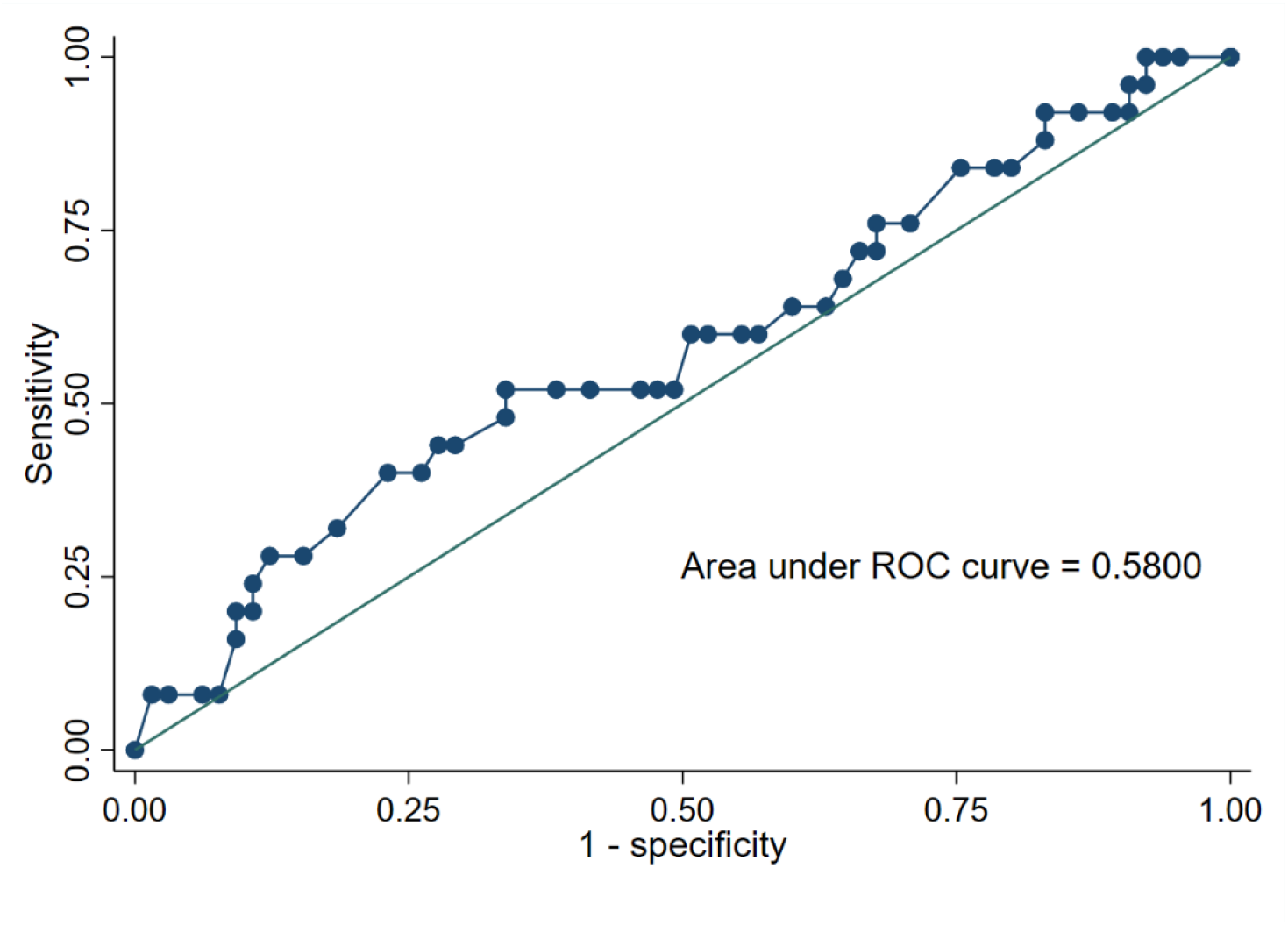
ROC curve for determining the CLIQ total score cut-off point distinguishing moderate and severe impact categories

#### Responsiveness to change

Forty participants had the CLIQ repeated at Day 90 following treatment. The median CLIQ score of this group at enrolment was 47.0 and 40.5 at Day 90. The difference in median CLIQ scores was significant (P-value= 0.037).

There was no significant difference in CLIQ scores of individuals who were deemed cured or improved at Day 90 by the treating physician. However, there was a significant difference (p = 0.044) in the CLIQ score of participants who rated their improvement as “better” at Day-90 compared to their CLIQ score at enrolment **(Figure 4**).

**Figure 4.**
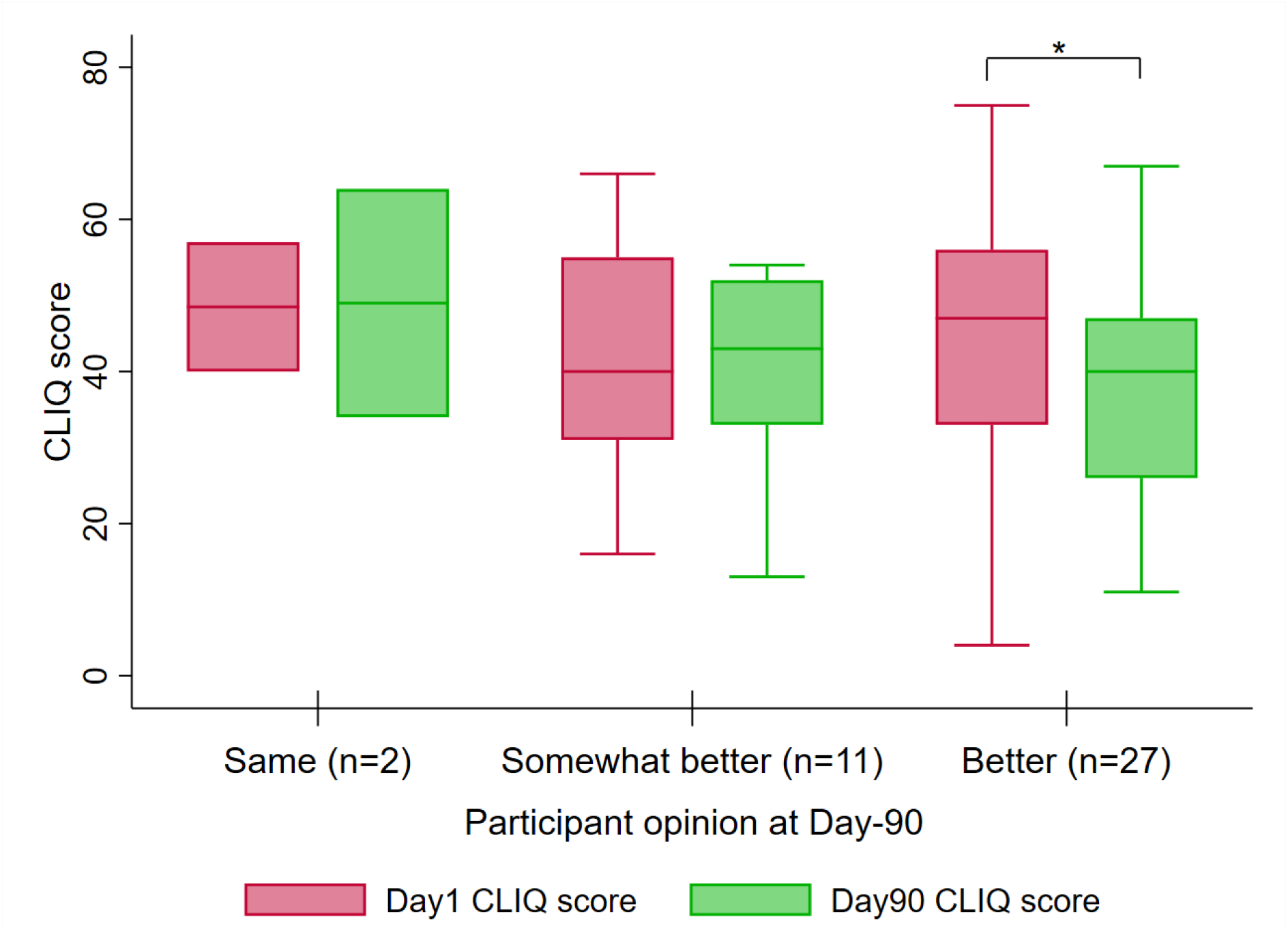
Boxplot of CLIQ Total Scores at Day 1 and Day 90, Stratified by participant opinion of their clinical outcome at Day 90. Note: * statistically significant difference (P value= 0.044)

#### Clinical Importance Difference (CID)

Participant anchor-based assessment of CID revealed that the mean change for those who reported themselves to be “better” was -7.3 (±18.5). Participants who felt “somewhat better” had a mean change in CLIQ score of -1.4 (±13.3). The difference in mean change in CLIQ scores between those who were “better” and those who were “unchanged” was 6.8. Using a distribution-based approach, the CID was 9.0 with a medium effect size (0.5).

## 4. Discussion

Disease-specific PROMs capture the unique experiences and perspectives of affected individuals providing valuable insights into HRQoL (22). In this study, the English version of CLIQ was translated into a culturally adapted, reliable and valid Afan Oromo version. Items 4 and 14 were adapted by adding a six-month recall period to clarify health expense reporting and introducing a “Not applicable” option for sexual difficulties. The content validity of the Afan Oromo CLIQ was consistent with the original Brazilian Portuguese CLIQ (9) and our recent Amharic translation (23). It displayed excellent internal consistency comparable to the original (9), as well as excellent inter-rater reliability similar to the Amharic version (23). Our findings support the original two-cluster structure of the CLIQ (9), with strong convergent and discriminant validity indicating that the Afan Oromo version accurately measures the intended HRQoL construct.

Afan Oromo CLIQ scores differentiate CL affected participants with different physician-determined severity and by lesion size which is consistent with more severe or larger lesions being associated with worse HRQoL measured using the Dermatology Life Quality Index (DLQI) in Ethiopia and other settings (24–26). We established CLIQ scores cut-off points to classify CLIQ scores as mild (<50) or moderate (50-54) or severe (>54) potentially offering a more objective approach to categorizing patient-reported impact. In contrast, a previous study (27) classified patient-reported impact by dividing participants into two groups based on the median score of clusters (low and high impact for cluster 1, and low and high satisfaction for cluster 2). Individuals with scores below the sample median were categorized as having lower impact, whereas those above the median were considered to have higher impact.

The Afan Oromo CLIQ demonstrated responsiveness to change assessed at Day 90 following treatment with participants’ who rated themselves “better” having significantly improved scores. This pattern is similar to findings from the Brazilian Portuguese (8) and Amharic versions (23), and another longitudinal study conducted in Brazil (28), where substantial reductions in CLIQ scores were observed following treatment, indicating that the instrument effectively captures meaningful clinical improvement over time.

We established a CID of 7 units, using anchor methods. This provides a meaningful benchmark for interpreting change over time, enabling clinicians and researchers to assess meaningful clinical change for affected individuals. The CID for the Amharic version of the CLIQ was 12, based on anchor-based analyses and supported by distribution-based estimates (23).

The availability of validated translations of the CLIQ in Afan Oromo and Amharic provide important tools for researchers assessing the burden of CL and the effectiveness of treatments in Ethiopia. Prior to these studies no validated disease specific measures of CL were available limiting quantitative assessment of CL from the perspective of affected individuals. However, given the different properties of the two language versions researchers must be cautious in combining CLIQ scores generated using the Amharic CLIQ and Afan Oromo CLIQ.

The limitations of our study include lack of a comparative assessment between the translated Afan Oromo CLIQ version and a validated Afan Oromo HRQoL tool, which was not available during our study. Another limitation is the small number of individuals who reported their clinical change as “same”, “somewhat worse”, and “worse” at Day 90, this limits an analysis of minimal clinically important difference (MCID), defined as the smallest change in a patient reported outcome that patients perceive as meaningful or clinically important (16). Although our sample size was adequate for reliability and validity analyses (29–31), a larger sample size would have been beneficial to strengthen the findings. The subjective nature of physician assessed CL severity and participant self-assessment responses may have introduced bias.

We have shown that the Afan Oromo CLIQ is a culturally appropriate, reliable, and valid instrument to measure the impact of CL in Afan Oromo-speaking adults. Further studies are needed to establish the MCID of the Afan Oromo CLIQ, adapt and tailor the CLIQ for children and adolescents.

## Data Availability

Data available on request.

## Statements & Declarations

### Ethics Approval

The study was approved by the National Ethics Review Committee of Ethiopia (ref no: 7/2-506/m259/35 dated 17.12.2021), the ALERT/AHRI Ethical Review Committees (ref no: PO/23/21 dated 15.07.2021), and the London School of Hygiene & Tropical Medicine Research Ethics Committee (ref no: 26421 dated 11.10.2021). Written informed consent was obtained from all participants.

### Consent for publication

Not applicable

### Availability of data and materials

Data available on request.

### Funding

The Skin Health Africa Research Programme (SHARP) is funded by the Research and Innovation for Global Health Transformation (RIGHT) Programme [Grant Reference Number NIHR200125] of the National Institute for Health and Care Research (NIHR) https://www.nihr.ac.uk/. The funder had no role in study design, data collection and analysis, decision to publish, or preparation of the manuscript.

Sponsor: London School of Hygiene & Tropical Medicine

FUNDERS: UK National Institute of Health Research Grant Ref: NIHR200125

### Author Contributions

Conceptualization: Stephen L. Walker, Endalamaw Gadisa, Michael Marks, and Saba Maria Lambert

Data Collection, Management and Curation: Debisa Eshatu Wendimu, Derese Bekele Daba, Mosisa Bekele Degefa, Amel Beshir Mohammed, Fewzia Shikur Mohammed, Shimelis Nigusie Doni, Belachew Hailu, Teklu Cherkose, Kibrome Tekleab, Tadele Molla, Tewodros Kaleb Kassa, Endale Hailu

Formal Analysis: Debisa Eshatu Wendimu

Funding acquisition: Stephen L. Walker, Endalamaw Gadisa, Michael Marks Saba Maria Lambert

Project administration: Eyerusalem Tesfaye Beyene

Software: Fikregabrail Abera, Tedros Nigusse Ferede, Galana Mamo

Supervision: Stephen L. Walker, Endalamaw Gadisa, Yohannes Hailemichael, Saba Maria Lambert, Sagni Chali Jira,

Writing – Original Draft: Debisa Eshatu Wendimu, Eyerusalem Tesfaye Beyene

Writing – Review & Editing: All Authors

## Acknowledgements

We wish to thank the individuals and communities for their participation in the work of the Skin Health Africa Research Programme.

We acknowledge our fellow leishmaniasis researchers, Professor Endi Lanza Galvão, Dr Ana Rabello and Dr Gláucia Cota, who developed the original CLIQ

## Notes

**Conflicts of Interest** None to declare

### Competing Interest Statement

The authors have declared no competing interest.

### Clinical Protocols

https://pubmed.ncbi.nlm.nih.gov/39839605/

### Funding Statement

Yes

